# A clinically integrated, frameless human Neuropixels workflow

**DOI:** 10.64898/2026.05.07.26351853

**Authors:** Hugo Layard Horsfall, Ahmed K Toma, Laurence Watkins, Harith Akram, Hani J Marcus, Ainsley Stewart, Jake Chatburn, Anne Vanhoestenberghe, Brian F Coughlin, Angelique C Paulk, Sydney S Cash, Marleen Welkenhuysen, Barundeb Dutta, Andreas T Schaefer, Mihaly Kollo, William Muirhead

## Abstract

High-density electrophysiological recording with Neuropixels probes enables single-unit resolution of human neural activity, but integration into routine clinical environments remains challenging and reported recordings have been limited to a few US centres. Here, we present a reproducible roadmap for human Neuropixels recordings under a nationally managed regulatory framework in the United Kingdom. Guided by the IDEAL Stage 2a (Development) framework, we established a frameless intraoperative workflow using manufacturer-sterilised probes and a commercially available, clinical-grade setup. We prospectively evaluated this workflow in six participants (mean age 62.5 years) undergoing elective ventriculoperitoneal shunt surgery, assessed across three predefined endpoints: clinical safety, procedural timing, and neural data yield. Iterative failure-mitigation cycles resolved key technical barriers, including neuronavigation interference and hardware instability. The workflow achieved zero research-related adverse events, maintained a 30-minute procedural extension, and progressively increased single-unit yield to 146 manually curated units.

## Introduction

The introduction of Neuropixels has driven a paradigm shift in systems neuroscience in animals, routinely enabling the simultaneous recording of hundreds of individual neurons across cortical layers with unprecedented resolution(Jun et al., 2017). Pioneering efforts in the United States have translated Neuropixels to humans and demonstrated initial safety and feasibility of high-density recordings during neurosurgery, with important neuroscientific insights relating to speech mechanisms(Chung et al., 2022; Chung et al., 2025; Coughlin et al., 2023; Leonard et al., 2024; Paulk et al., 2022). However, Neuropixels in humans remains technically challenging. The study setup requires capital investment, stringent regulatory and clinical oversight, and a specialist interdisciplinary team with a wide range of skills ranging from clinical neurosurgery to computational neuroscience and engineering. Participant recruitment is another consideration. These complex factors are a barrier to the broader adoption of Neuropixels in humans in different environments. For Neuropixels to reach its full potential for exploratory and translational neuroscience (Lee et al., 2024), high-density human electrophysiology must transition toward a robust, reliable workflow that integrates with routine operative workflows and broader systems internationally. A global consortium capable of high-density human neurophysiology provides a mechanism for the development of standardised neural data quality management and metrics via cross-validation, and an open-science approach to human discovery.

Neuropixels are a complex research tool used during surgical intervention. Evaluation and assessment of techniques and complex interventions is multidimensional and challenging. The IDEAL (Idea, Development, Exploration, Assessment, and Long-term study) framework provides structured evaluation across different life cycle stages, and offers bespoke metrics for different stages(McCulloch et al., 2009). IDEAL considers four domains: the device, clinicians, the patient, and wider systems during evaluation. Whilst IDEAL is primarily used to evaluate clinical techniques and devices, the core philosophy can be applied to research tools such as Neuropixels. The IDEAL principles can be applied to structure the description and integration to clinical workflow. Already for Neuropixels, IDEAL Stage 0 (Preclinical) rodent and non-human primate research validated the safety and recording efficacy required for “bench-to-bedside”, and US-led first-in-human studies demonstrated safety for first-in-human IDEAL Stage 1 (Idea) (Marcus et al., 2022; McCulloch et al., 2009, Paulk et al., 2022; Chung et al., 2022). Adoption and scaling of intraoperative human high-density electrophysiology requires Stage 2a (Development), whereby the learning curve of a new procedure focuses on the iterative refinement to the technique, the stabilisation of surgical workflow, and the formalisation of safety protocols (Diez del Val et al., 2015). Recently applied to evaluate high-stakes surgical innovations such as surgical robotics (Marcus et al., 2024), the IDEAL framework encourages an approach that considers regulatory hurdles, clinical constraints, and safety and human factors. Using the IDEAL framework, we report iterations towards a maturing, clinically integrated human Neuropixels workflow which is reproducible and capable of delivering high-fidelity neural data.

We present the regulatory considerations during study setup for non-approved devices like Neuropixels to be used in humans in a non-US environment. We describe iterative technical developments beyond research tools to clinical, hospital-approved equipment sterilised in routine workflow, within a nationalised healthcare and regulatory framework of the National Health Service (NHS, UK). We introduce a non-head-fixed Neuropixels insertion method which allows different patient populations to participate in high-density electrophysiology research. Finally, we report failure-mitigation loops required to achieve stable recordings within the IDEAL 2a framework. In doing so, we provide a structure for the global adoption of Neuropixels in humans to enable centres to contribute to the collective understanding of the human brain.

## Results

### Establishing a national regulatory pathway for Neuropixels in humans

The initial barrier to establishing a Neuropixels workflow was navigating the regulatory constraints associated with non-certified devices. Neuropixels probes are currently designated for research use only without CE-, or UKCA marking and in the US, does not have Food and Drug Administration (FDA) clearance for human clinical use. The US groups navigated this through local Institutional Review Board (IRB) approvals to utilise the probes as modified research tools (Coughlin et al., 2023). Local IRB approvals are non-trivial. However, translating this technology into the centralised regulatory framework of the United Kingdom’s National Health Service required a different approach to ensure reproducibility and compliance.

To establish a national regulatory roadmap, we consulted the Medicines and Healthcare products Regulatory Agency (MHRA), and utilised the MHRA online Flow Chart for Clinical Investigations under UK Medical Device Regulations (UK MDR; Supplementary Figure 1). Although Neuropixels are used in humans and do not have CE- or UKCA marking as a medical device, they are not being used for medical purposes but as a research tool. Therefore, this does not require notification to the MHRA, and through subsequent direct correspondence, it was confirmed that formal notification was not required.

This regulatory outcome was a critical first step in our workflow development. However, it shifted the burden of safety and technical validation from national MHRA-led oversight to rigorous assessment by local biomedical engineering and governance teams at the participating hospital. Consequently, this required our team to integrate equipment with appropriate CE and UKCA certification (such as a medical-grade micromanipulator) to satisfy local safety requirements and routine clinical workflows, including hospital sterilisation protocols, all within the context of robust ethical safeguards. This pathway ultimately provided the necessary institutional clearance to commence the IDEAL Stage 2a technical maturation.

### Towards clinically integrated high-density human neurophysiology

We prospectively developed a workflow evaluated across IDEAL’s four domains: device, clinician, patient, and system. The workflow maturation was evaluated against three primary endpoints: Endpoint 1 (E1): Notifiable Safety Events (NSE) including shank fractures; Endpoint 2 (E2): duration (research extension and recording time); and Endpoint 3 (E3): yield (successful recording and putative unit yield).

Six participants were prospectively recruited at a quaternary academic neurosciences centre (n=6; 1 male, 5 female; mean age 62.5 years; range 41-79 years), provided written informed consent, and underwent elective ventriculoperitoneal shunt (VPS) insertion (Table 1; Figure 1). This cohort represents an older patient population than is typically reported in the human Neuropixels literature. Clinical safety remained robust across all cases (E1), with no research-related intraoperative events, zero shank fractures, and no adverse events reported at the 30-day follow-up (Table 1).

**Figure 1.**
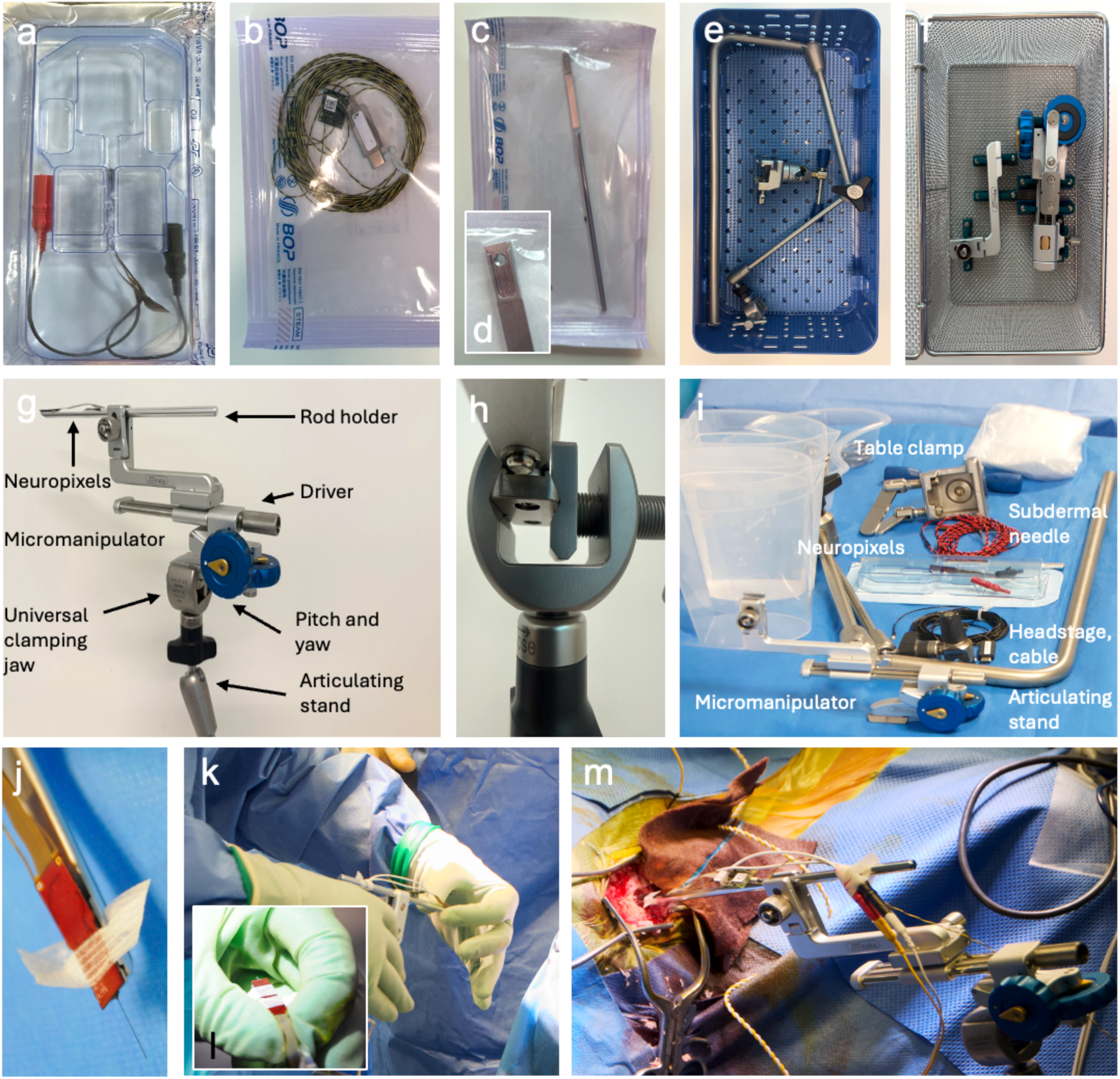
Clinically integrated, frameless workflow for intraoperative human Neuropixels recordings. (a) Neuropixels 1.0 NHP short-S probe in primary blister packaging following manufacturer-led Ethylene Oxide (EtO) sterilisation, with pre-soldered grounding leads. (b-d) Manufacturer-sterilised headstage (HS_1000) connection cable (CBL_1000-S), and rod holder (HOLDER_1000_C-S). (e) Karl Storz articulating stand in sterilisation tray. (f) Mitaka micromanipulator in sterilisation tray. (g) Assembled micromanipulator–endoscope holder complex with labelled components, providing sub-millimetre control in pitch, yaw, and translational advancement. (h) Close-up of the Karl Storz distal clamping jaw (introduced at v1.3). (i) Complete sterile intraoperative equipment preparation with components labelled. (j) Neuropixels probe mounted on rod holder with steristrip “wings” for safer handling. (k) Intraoperative transfer of the rod holder–Neuropixels complex to the operative field. Inset (l): recommended “pincer grip” handling technique (Coughlin et al., 2023). (m) Assembled v2.0 configuration during recording in SU007, showing frameless insertion using a standard horseshoe headrest aligned orthogonally to the right parieto-occipital cortex (Brodmann area 39).

**Table 1.** Cohort characteristics with intraoperative Neuropixels Endpoints (E). Summary of participant demographics, diagnosis, operation, and Neuropixels Endpoints (E1-3). Six participants undergoing elective ventriculoperitoneal shunt insertion were prospectively recruited. All recordings were performed in the right parieto-occipital cortex (estimated angular gyrus/Brodmann area 39*). Recording duration reflects the Neuropixels recording within the 30-minute operative extension, with the optimal recording to last for the full permitted 15 minutes. Putative unit yield reflects spike-sorted single units following automated spike sorting and manual curation. No Neuropixels shank fractures, intracranial haemorrhages, infections, or research-related adverse events were observed intraoperatively or at 30-day follow-up. One participant (SU004) consented but did not undergo recording due to postponement of surgery for clinical reasons and was excluded from analysis. NPH: normal pressure hydrocephalus; LOVA: longstanding overt ventriculomegaly of adults; PO cortex: parieto-occipital cortex; VP shunt: ventriculoperitoneal shunt. E1 Fracture describes shank fracture and NSE: Notifiable Safety Event either intraoperatively or at 30-day follow-up; Yield: putative units following data preprocessing, motion correction, spike sorting and manual curation. *Age range to comply with preprint conditions.

| Case | Age* | Sex | Diagnosis | Side | Location* | Operation | Neuropixels inserted | E1: NSE |  | E2: Duration, mins |  | E3: Data, units |  |
| --- | --- | --- | --- | --- | --- | --- | --- | --- | --- | --- | --- | --- | --- |
|  |  |  |  |  |  |  |  | Fracture | Other | Research | Record | Data | Putative units |
| SU001 | 76-80 | M | NPH | Right | PO cortex | VP shunt | Y | N | N | 28 | 7 | Y | 25 |
| SU002 | 76-80 | F | NPH | Right | PO cortex | VP shunt | Y | N | N | 20 | - | N | N |
| SU003 | 41-45 | F | Early midlife hydrocephalus | Right | PO cortex | VP shunt | Y | N | N | 20 | - | N | N |
| SU005 | 46-50 | F | Late midlife hydrocephalus | Right | PO cortex | VP shunt | Y | N | N | 27 | 10 | Y | 70 |
| SU006 | 51-55 | F | Late midlife hydrocephalus | Right | PO cortex | VP shunt | Y | N | N | 29 | 2 | Y | Unable to sort |
| SU007 | 76-80 | F | NPH | Right | PO cortex | VP shunt | Y | N | N | 24 | 15 | Y | 146 |

The iterative implementation of failure-mitigation loops was central to the maturation of the workflow (Figure 2). Technical challenges identified in early cases (v1.0–v1.2) specifically identified neuronavigation systems as the primary source of noise in SU002 and SU003. Process maturation continued through v1.4, where hardware instability with the initial PXIe acquisition chassis drove the refinement of our hardware protocols. This development process culminated in the v2.0 stabilised workflow utilised for SU007, which successfully achieved the primary data endpoint (E3) with 146 putative units recorded during a full 15-minute session (Figure 3). These longitudinal results provide the prospective technical and procedural data for the standardised domains discussed below (Table 2; Supplementary Table 2).

**Figure 2.**
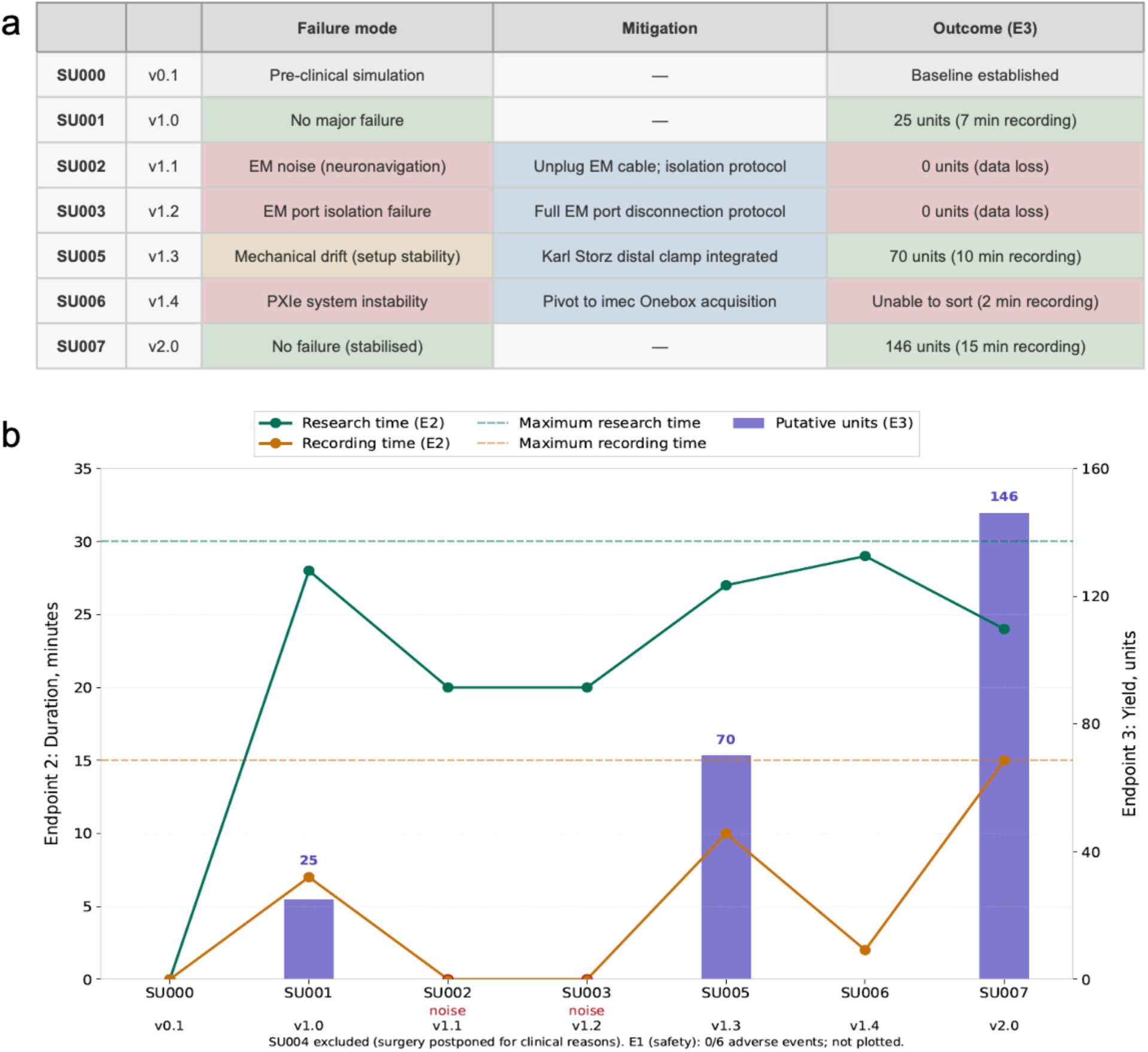
Iterative development and failure-mitigation across Neuropixels recordings (IDEAL Stage 2a). (a) Prospective workflow maturation across consecutive cases (SU000–SU007) documenting failure modes, mitigations, and outcomes. SU000 was preclinical simulation. SU004 was excluded as surgery was postponed for clinical reasons. (b) Iterative improvement in duration (E2) and putative units (E3) endpoints. Endpoint 2 (left y-axis): total research extension time (green) and active recording duration (orange) per case. Dashed lines indicate the maximum permitted 30-minute research extension and 15-minute recording window. Endpoint 3 (right y-axis, purple bars): putative single-unit yield following automated spike sorting and manual curation. Endpoint 1 (safety) is not plotted as there were zero shank fractures or adverse events across the cohort. Version annotations (v0.1–v2.0) denote iterative workflow changes, culminating in a stabilised pipeline (v2.0) achieving 146 units within the full 15-minute recording window. E1, Endpoint 1 (safety); E2, Endpoint 2 (duration); E3, Endpoint 3 (yield); EM, electromagnetic; v, version.

**Figure 3.**
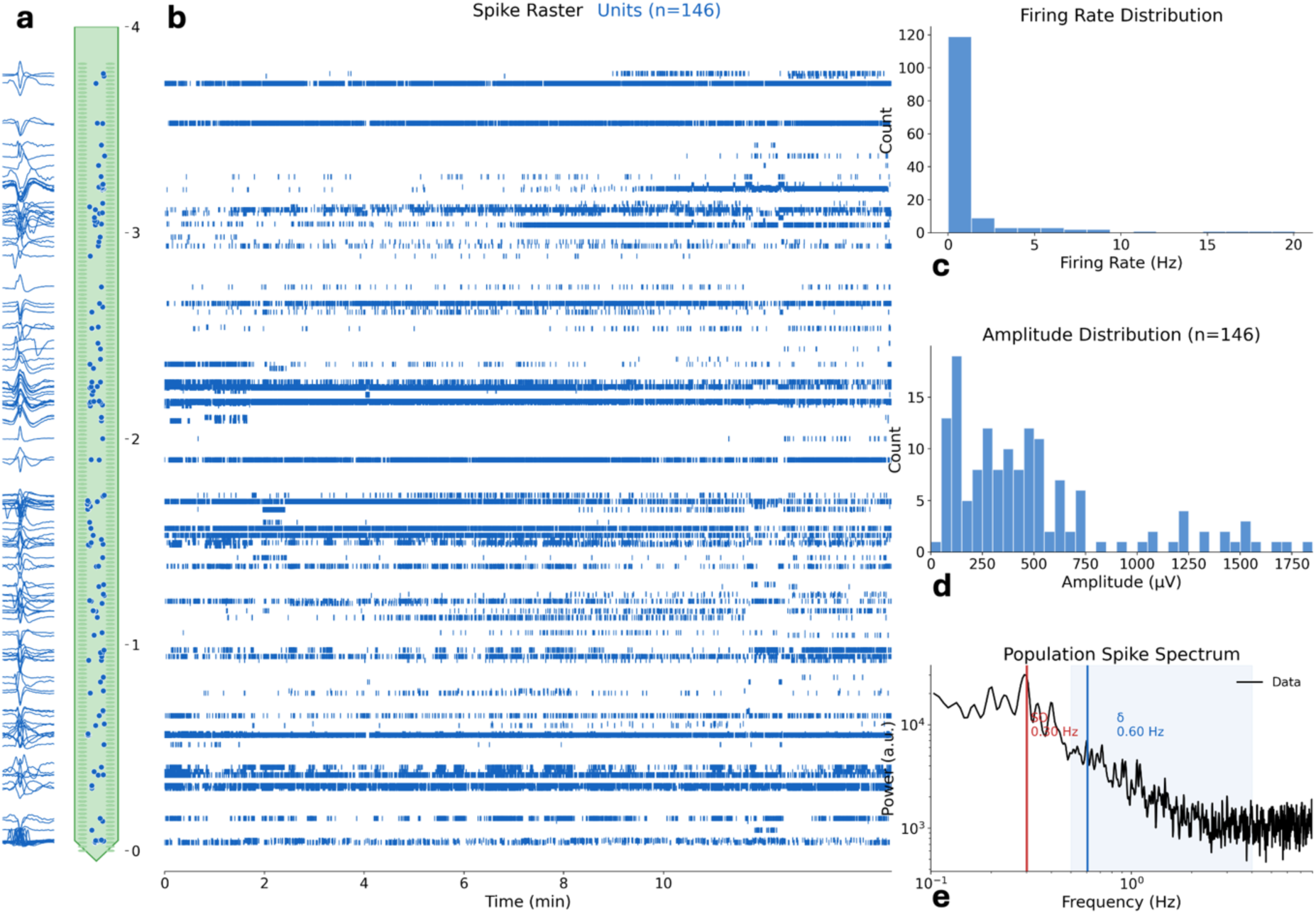
Representative single-unit neural data from the stabilised workflow (SU007, v2.0). **(a)** Mean waveforms of individual units displayed at channel locations along the Neuropixels shank with approximate cortical depth span of recorded units. Blue traces show mean waveform morphology. **(b)** Spike raster plot for all 146 manually curated single units across the full ∼15-minute recording window. Each row represents one unit, ordered by depth (probe tip at bottom). Depth (mm) is indicated on the left axis relative to the cortical surface. **(c)** Distribution of mean firing rates across all 146 units. The majority of units fired below 5 Hz (mean 1.35 ± 3.35 Hz), consistent with sparse cortical activity under general anaesthesia. **(d)** Distribution of spike amplitudes across all 146 units (mean 262 ± 242 µV). **(e)** Population spike spectrum (power spectral density of the aggregate spike train). The spectrum reveals peaks at approximately 0.34 Hz and 0.60 Hz, possibly consistent with slow oscillatory modulation of population firing characteristic of propofol-induced anaesthesia.

**Table 2.** Taxonomy of a clinically integrated human Neuropixels pipeline. The perioperative workflow is mapped against the four interdependent domains of the IDEAL framework (Device, Clinician, Patient, and System). This matrix provides a chronological blueprint of the regulatory, technical, and human factors required to safely translate high-density electrophysiology into a routine surgical environment. By structuring the pipeline across pre-operative, operative, and post-operative, this taxonomy serves as a reproducible checklist for multi-centre adoption under the IDEAL Stage 2a development paradigm.

| Phase | Device | Clinician | Patient | System |
| --- | --- | --- | --- | --- |
| **Pre-operative: setup and planning** | Source clinical-grade, CE/UKCA/FDA marked hardware (eg micromanipulator)<br>Establish sterilisation pathways (eg manufacturer-led EtO)<br>Validate acquisition software and hardware | Select appropriate patient cohorts and design workflow<br>Conduct pre-clinical simulations with silicon brain phantoms<br>Train non-study surgeons and scrub teams | Conduct Patient & Public Involvement (PPI) sessions<br>Informed consent<br>Communicate transparently regarding risk and lack of direct benefit | Secure regulatory exemptions (eg MHRA research tool / FDA IRB waiver)<br>Obtain ethical approvals<br>Establish Data Management Plan<br>Ensure local biomedical engineering compliance and risk assessment |
| **Operative: Neuropixels recording** | Integrate equipment into the sterile field (eg Onebox, sterile draping)<br>Achieve electrical isolation and grounding<br>Perform intraoperative troubleshooting/reboots<br>Obtain neural recording | Perform frameless insertion and orthogonal alignment<br>Avoid macroscopic cortical vessels during pial penetration<br>Adhere to operative extension caps (eg maximum 30 minutes)<br>Monitor physiological stability under general anaesthesia | Participant intraoperative task as appropriate | Conduct WHO-style pre-operative briefing<br>Define interdisciplinary role allocation<br>Manage theatre ergonomics and human factors<br>Perform theatre de-noising checklist |
| **Post-operative: analysis** | Inspect explanted probe for shank fracture<br>Ensure safe sharps disposal and device accountability<br>Hardware cleaning and sterilisation | Conduct formal surgical and team debriefs for iterative improvement<br>Review of participant post-operatively and 30 day follow up for research related adverse events (eg haematoma, infection)<br>Design failure-mitigation loops for the next case | Inform patient about outcome of study as appropriate | Sample management if appropriate<br>Manage data governance and storage<br>Transfer data to computational pipelines and analysis |

### Device: transitioning from research to routine neurosurgical equipment

The maturation of the Neuropixels workflow relied upon leveraging existing innovations in thickened shanks and sharpened tips developed by international colleagues to reduce shank fracture during setup and pial penetration. While early literature reported fractures or mechanical stress (bending) during insertion, our immediate adoption of the Neuropixels NHP 1.0-s resulted in no shank fractures (E1) across our cohort. Manufacturer-led sterilisation of the probe with pre-soldered grounding cables, headstage and connection cables via Ethylene Oxide (EtO) validated to ISO standards, delivered in familiar surgical-grade primary and secondary packaging, contributed to efficient equipment workflow and satisfied local requirements for regulatory considerations of device sterility. The result is an end-to-end clinically integrated system that did not require bespoke Neuropixels sterilisation trays or in-house EtO sterilisation pathways, and negated technically challenging sterile ultrasound sleeve envelopment of the research system (Figure 1). This also facilitated using the commercially available, manufacturer-sterilised stereotactic rod holders, eliminating improvised research techniques in the operating theatre which can be difficult to implement, both technically and from a regulatory perspective.

Our neural data acquisition system used an NHS-approved cart featuring a theatre-appropriate form factor, medical-grade power isolation, and a clinical-grade keyboard and mouse familiar to all operative staff. Initial cases used the National Instrument PXIe box in accordance with established literature. However, this research tool has a large footprint and mandates time-consuming system reboots during intraoperative debugging. We subsequently acquired the imec Onebox following its release and validated its safety in collaboration with our local biomedical and physics engineering team. During the SU006 case (v1.4), a technical failure where the computer failed to recognise the headstage via the PXIe box resulted in a truncated recording of only ∼2 minutes due to the multiple full-system reboots required which takes 3-5minutes per reboot, versus the Onebox which is ∼20 seconds. We subsequently integrated the Onebox for SU007, which contributed to a stabilised workflow achieving our highest yield (146 units) within the 15-minute recording window, even including two reboots required to debug the system intraoperatively (Supplementary Table 1). The Onebox is clinically ready due to its small form factor and independent reboot capability, also allowing the Neuropixels setup to be performed entirely within OpenEphys.

### Clinician: integrating standardised equipment

We leveraged existing clinical equipment compliant with regulatory and sterilisation standards to reduce procedural friction, including a Karl Storz endoscope holder connected to the surgical bed and a Mitaka micromanipulator (Figure 1). An attachment (point setter universal attachment, see Methods) from the micromanipulator attaches to the manufacturer-sterilised rod holders, which allows a stable Neuropixels setup without the need for non-regulated research tools. The micromanipulator provides sub-millimetre control in pitch and yaw (angles, degrees), and advancement (displacement, mm), ensuring an orthogonal and stable insertion into the cortex. A specific clamp attachment for the Karl Storz endoscope holder was identified and iteratively integrated into the setup for SU005 (v1.3) (Figure 1h). The clamp had better grip on the micromanipulator and increased mechanical stability to reduce lateral movement during insertion and increasing unit yield (E3: SU001 25 units without clamp attachment vs SU005 70 units with clamp attachment). A custom sterilisation tray was made for the micromanipulator, and the manufacturer-sourced sterilisation tray was used for the endoscope holder. The endoscope holder and micromanipulator were sterilised within the routine hospital workflow.

This clinically integrated hardware facilitated participants who were not head-fixed in a Mayfield clamp or stereotactic frame to participate in research using a standard operating theatre horseshoe headrest. This “frameless” approach enabled high-density recordings in an older, previously not recorded from, patient population.

Familiarity with this clinical-grade equipment amongst surgeons and scrub teams reduced the cognitive burden of a new research procedure in a time-pressured operative environment. The intuitive nature of the endoscope holder and micromanipulator allowed non-study surgeons to microscopically orient the probe, avoid cortical vessels, and optimise insertion trajectory. Such precision is vital for maximising unit yield and minimising lateral mechanical forces on the shank, thereby further reducing the risk of fracture. There were no failure modes related to the clinical equipment introduced to the workflow, either from a procedural (i.e. sterilisation) or technical (i.e. unable to get orthogonal insertion) perspective.

### Patient: communicating a novel, high-complexity intervention with no direct benefit

The ethics of basic science research with no direct patient benefit mandate high levels of transparency regarding risk and uncertainty. Initial safety data and proof-of-concept guidance from US collaborators informed our patient-facing communication strategies. A Patient and Public Involvement session was conducted locally to explore acceptability of the Neuropixels research, particularly related to operative time extension and risk (Piper et al., 2025). This was important for developing patient focussed patient information sheets and informed consent processes.

Our protocol mandated a period of at least 24 hours between the initial provision of information and the formal consent discussion to ensure adequate time for the participant to consider the information provided. Remote consent via telephone or video conference was successful, often with family members present to ensure all relevant parties understood the research objectives and safety profile. We emphasised that participation offered no direct individual benefit and that choosing not to participate would not affect their routine clinical care.

To evaluate the real-world acceptability, we tracked recruitment metrics (Figure 4). Of 12 patients formally screened, 10 met the inclusion and exclusion criteria. Seven participants provided informed consent, representing a high uptake rate in an older population with comorbidities (successful recruitment: 70%). Three eligible patients declined:

- One patient (70’s, male) expressed concern that the 30-minute operative extension might exacerbate pre-existing renal disease.
- One patient (20’s, female) with a complex medical history preferred to avoid additional research interventions or testing beyond their routine care.
- One patient (80’s, male) declined due to the requirement for 30-day clinical follow-up and perceived risks regarding probe shank fracture.

**Figure 4.**
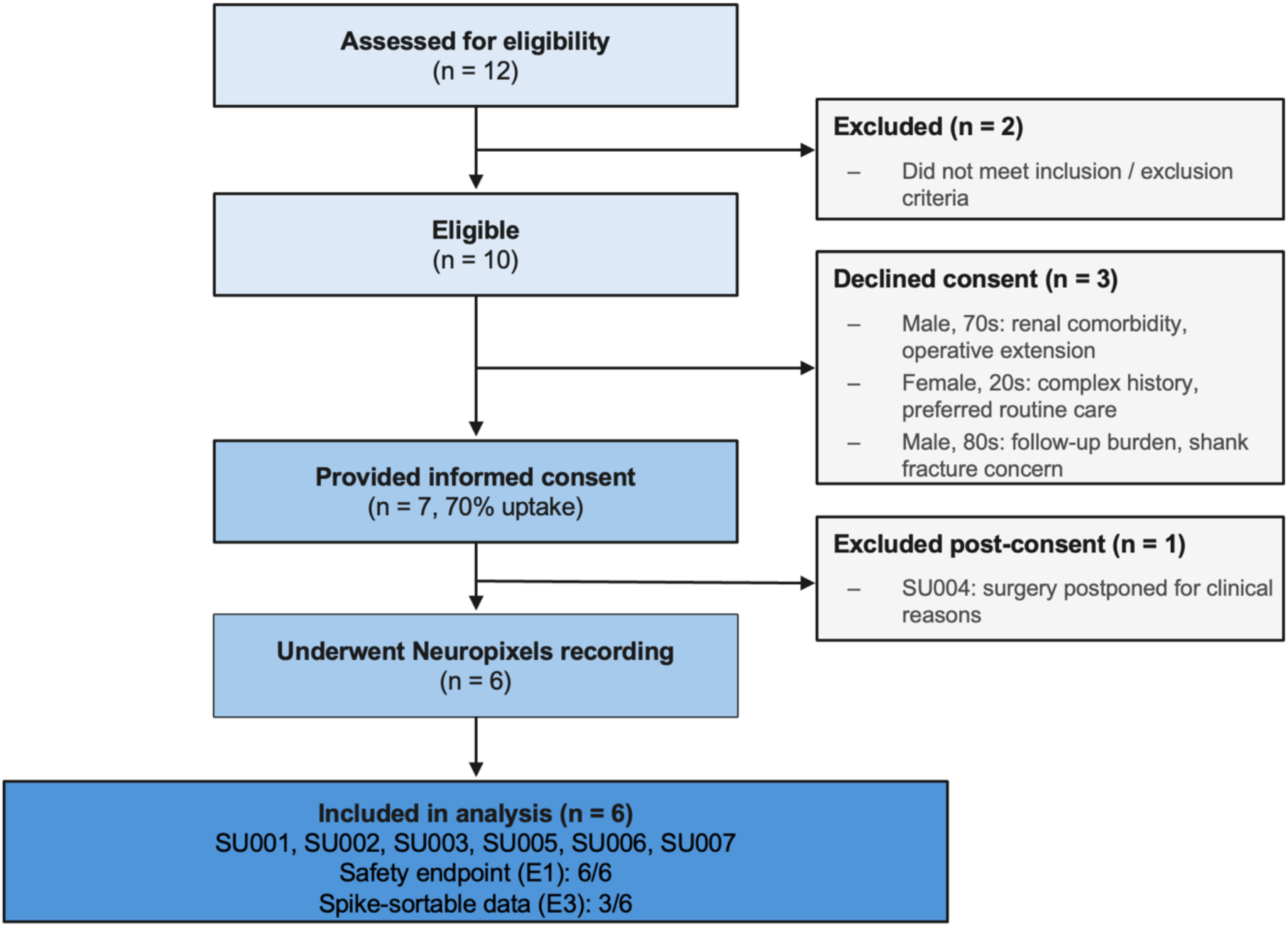
Participant recruitment. Of 12 patients screened, 2 were ineligible and 3 declined consent, leading to 7 consented participants (70% uptake). One consented participant (SU004) was excluded after surgery was postponed for unrelated clinical reasons. Six participants underwent Neuropixels recording and were included in analysis, with the clinical safety endpoint (E1) met in all cases and spike-sortable neural data (E3) obtained in three.

Ultimately, six participants underwent recording, as one participant (SU004) was excluded post-consent after their surgery was postponed for clinical reasons. The successful recruitment of an older cohort (mean age 62.5 years; range 41-79 years), including three 79-year-old participants, highlights Neuropixels recordings are acceptable to patients, with transparent communication regarding the nature of basic science research and the 30-minute operative extension for research essential.

### Systems: safety, training and human factors in a Neuropixels workflo**<u>w</u>**

Safety remained a primary concern throughout the design and implementation of our study. We monitored Neuropixels-related complications (E1) both intraoperatively and at a 30-day follow-up, with no adverse events recorded across the cohort. An independent clinician reviewer was recruited to classify the expectedness of any potential complications, and an internal governance procedure was established to discuss events at departmental mortality and morbidity meetings. A pilot phase at n=5 was integrated for formal safety review by the Trial Steering Committee.

Training was delivered through three primary streams (simulation; wider team; neurosurgeons) to de-risk the intraoperative procedure and ensure the procedural timing endpoint (E2) was maintained within the 30-minute research cap. The time constraint aimed to (1) minimise risk to participants by anaesthetic extension; (2) minimise extension to routine operative lists; (3) maximise the neural recording window. Comprehensive pre-clinical simulation to practice the Neuropixels and theatre equipment setup (Supplementary Figure 2), and repeated insertions contributed to technique optimisation and awareness of high-risk critical steps for study success, such as the transfer of Neuropixels from setup table to the micromanipulator, or surgical instruments in the operative field near the Neuropixels shank and shank retrieval should it occur (Supplementary Figure 3). Further, targeted training through presentations and hands-on training with the theatre and scrub team, and the anaesthetic team, familiarised staff with the overall aim of the Neuropixels research, specific roles intraoperatively, strategies for noise mitigation, and troubleshooting strategies. Finally, a dedicated 5-point training program that covered the handling, workflow, insertion, recording, and removal of Neuropixels was delivered to neurosurgeons to improve understanding, confidence, and reduce the risk of any complications and contributed to the reassuring safety profile from the first 6 cases.

In addition to the simulation and training, we introduced World Health Organisation-style surgical checklists (Appendix 1). This included a pre-surgery checklist, performed by the research team during the surgical team brief in the morning. Here, the research team re-introduced themselves, assigned and clarified roles, the purpose of the research, and checked patient safety data such as confirmed ongoing consent, inclusion criteria met, pregnancy status, ASA grade, and, if appropriate, which behavioural tasks may be performed. At this point, we also re-iterated the importance of theatre electrical de-noising and steps to take to ensure the high quality of the recording. The signed consent form was also re-checked during WHO “Time Out” as surgery began. Another checklist was intraoperative, prior to the Neuropixels insertion, where the electrical equipment was sequentially turned off with closed-loop communication between the research and clinical team to ensure the action was complete. Lastly, taking inspiration from the MGH team (Coughlin et al., 2023), we designed a granular operative workflow that spanned pre-operative, intraoperative, post-operative which included troubleshooting escalation strategies.

This multifactorial approach towards Neuropixels, and the subsequent clinical learning curve resulted in a granular, iterative workflow maturation (v0.1–v2.0). This system proved essential following the failure modes identified in SU002 and SU003, where electromagnetic interference from neuronavigation led to data loss (E3). In response, we developed and implemented a theatre de-noising checklist to ensure full EM isolation prior to insertion. Finally, the research team maintained an open culture that utilised post-operative debriefs to identify hardware instabilities, such as those encountered in SU006, leading to the successful pivot to the Onebox acquisition system and workflow stabilisation.

## Discussion

### A systems approach to standardising high-density neurophysiology

The translation of Neuropixels from laboratory to viable clinical research instrument necessitates advancement from research technical adaptations to a formalised systems-engineering approach. The IDEAL framework (Idea, Development, Exploration, Assessment, Long-term study) is applied to new surgical interventions and establishes the complex evaluation metrics across four domains: device, clinical, patient, and systems. Recently, IDEAL has been applied to evaluate high-stakes surgical innovations such as surgical robotics (Marcus et al., 2024). Although the use of Neuropixels in humans currently is for exploratory science rather than diagnostics or therapeutics, we believe there are valuable lessons that can be adopted by groups working with Neuropixels in humans, such as regulatory considerations, local requirements, clinical aspects and the learning curve, and safety and human factors inherent in neurosurgery. As such, we have prepared a table to support this (Table 2). This approach has facilitated the safe translation of Neuropixels into a new regulatory and clinical environment outside of the US.

Since their introduction for simultaneous high-density single-unit recordings in animals in 2017(Jun et al., 2017), Neuropixels have been adopted by thousands of neuroscientists worldwide, aligning with the IDEAL framework (Stage 0; preclinical). Pioneering US studies(Chung et al., 2022; Paulk et al., 2022) established the initial feasibility of recording in humans (IDEAL Stage 1; Idea) and resulted in an important technical evolution: the development of probes with increased shank thickness and a sharpened tip to facilitate easier pial penetration. This evolution was vital to reduce probe fracture – the primary safety concern for surgical teams. Our study builds on demonstrated initial feasibility to IDEAL Stage 2a (Development), the standardisation and maturity of the workflow for broader adoption.

Using an international framework like IDEAL provides a common language for the human neurophysiology field. It allows research groups to report iterative improvements – such as the recent technical innovation using 3D-printed tools and 45 mm long Neuropixels shanks (Daril E. Brown II et al., 2026) – within a standardised lexicon.

The progression toward IDEAL Stage 2b (Exploration) necessitates a shift from technical maturation to the multi-centre evaluation of procedural techniques and data consistency. Future efforts must systematically investigate how variations in surgical technique – such as comparing slow versus fast Neuropixels insertion rates – impact both mechanical safety (E1) and subsequent unit yield (E3). This phase will move beyond individual workflow stabilisation toward the formalisation of data quality and procedural benchmarks. Building upon metrics established by Chung and colleagues (Chung et al., 2022), the field must adopt standardised quality management systems for neural data alongside rigorous procedural benchmarks, including adverse events (E1), total operative extension time (E2), and recording yield (E3). By formalising these metrics within an open-science framework, the global human neurophysiology community can move away from ad-hoc reporting and towards a data-driven framework that balances high-fidelity scientific discovery with the uncompromising requirements of surgical safety and theatre efficiency (Chiong et al., 2018).

### Safety and utility of frameless Neuropixels in general anaesthesia

Safety remains a critical factor in the adoption of new technologies, particularly implantable devices to high-risk regions such as the human brain. Although safety had been demonstrated by our US colleagues(Chung et al., 2022; Chung et al., 2025; Paulk et al., 2022), safety continued to be our primary concern. Technical advancements in the probe, as discussed above, were critical for reducing shank fractures. We adopted a rigorous internal culture of simulation and interdisciplinary human factors training. Our study in the context of existing US literature provides evidence that Neuropixels can be safe even in new centres with different conditions and considerations. Importantly, the US groups’ willingness to collaborate and share early-stage learning experiences allowed us to bypass major pitfalls(Coughlin et al., 2023). While our cohort is small (n=6) compared to recent large-scale reports such as the 56-patient series from UCSF(Chung et al., 2025), it serves as a high-fidelity demonstration of how key components of the IDEAL framework can stabilise a complex, challenging technique towards a shorter learning curve and no complications.

The stability of our recording environment was achieved via clinical workflow utilising clinically integrated, surgical-grade equipment. The use of the Karl Storz endoscope holder and Mitaka micromanipulator provided multiple degrees of freedom and sub-millimetre precision for the insertion trajectory. Crucially, this allowed for perpendicular insertion relative to the cortex, maximising the quality of the laminar recording while minimising lateral movement that could lead to tissue trauma or probe fracture. Importantly, our frameless approach expands the patient populations eligible for participation. While traditional frame-based or head-fixed (Mayfield) setups are dominant in the literature for epilepsy or tumour resection, they are incompatible for patients undergoing procedures like shunt surgery. This also expands the age range beyond 30-50 years in the literature, and this study provides further evidence that a 30-minute extension to surgery is safe, and even in older patients with more comorbidities, Neuropixels is - so far - safe, feasible, and acceptable to patients.

Non-head-fixed Neuropixels procedures are performed under general anaesthesia (GA). This reduces the risk of shank fracture whereby a rigid probe connects to the bed in the context of an awake freely moving human. Therefore, scientific hypotheses differ versus an awake cohort. Numerous neuroscience dogmas derived from animal models have yet to be fully translated or explored in humans, and the unconscious brain offers a unique environment for such discovery. A recent preprint describes a Neuropixels technique to record unconscious learning in the human hippocampus during surgery(Katlowitz et al., 2025), and highlights that Neuropixels may be able to resolve complex circuit dynamics even outside of awake, task-based paradigms(Leonard et al., 2024). By expanding the patient cohorts and surgical contexts for Neuropixels, we provide a broader context in which to apply Neuropixels with the ultimate goal of developing a deeper understanding of human neurophysiology.

### Regulatory stewardship and the convergence of human neuroscience

The successful establishment of our Neuropixels workflow within a nationalised healthcare and regulatory system (NHS; MHRA) provides a roadmap for other centres. By navigating the MHRA regulatory exemption for non-commercial basic science research using a research tool, we demonstrate that high-stakes neurotechnology can be integrated into centralised healthcare infrastructures outside of local IRB models traditionally seen in the US. This pathway approach requires cross-institutional collaboration and support from local biomedical engineering and physics departments to ensure hardware safety and rigorous electrical denoising. However, the value of this effort extends beyond the technical yield of a single study.

We are in an era of unprecedented convergence of research modalities that promises to redefine our understanding of the human brain. As discussed by Lee et al., the integration of large-scale single-unit neurophysiology with other next-generation technologies such as single-cell RNA sequencing, spatial transcriptomics, and ex vivo tissue culture represents the next frontier of human neuroscience(Lee et al., 2024). In this paradigm, Neuropixels recordings provide the functional, circuit-level context for the underlying biological ground truth revealed by multi-modal tissue analysis workflow. By linking electrical signatures such as burstiness or laminar-specific neurons to their specific transcriptomic identities, we can move from descriptive reports of brain activity toward a mechanistic understanding of how cell-type-specific failures drive circuit dysfunction. This may support the development of new approaches to earlier or improved diagnostics and personalised therapeutics.

Key to Neuropixels research are the participants, whose contribution to basic science has no direct personal benefit, whilst increasing their exposure to real-world risk. Participant involvement was central to our protocol development, including consultation through Patient and Public Involvement sessions on acceptability towards Neuropixels, the research, and the length of operation extension. For Neuropixels or other intracranial recording devices, as newer tools emerge with increasing channel counts, longer probes, and converged “omics” workflows, the requirement for a safe, robust, and reproducible translational framework is essential. Such a framework and common language enable researchers and clinicians to demonstrate gradual iterations over time, moving from animal, first-in-human, to scaling of technology in a step-wise fashion that is transparent, and where safety and feasibility are the primary concerns. Once safety and feasibility have been demonstrated, it facilitates the opportunity to focus on the cellular underpinnings of the brain. Therefore, clinical frameworks like IDEAL applied to translational neuroscience ensure that device innovation is transparent, that risks are informed and carefully managed, and that learning curves and iterations are documented with scientific accuracy. This should reassure participants, as well as key stakeholders such as industry, clinicians, hospital management, and the regulators.

### Study Limitations

Despite the successful introduction of a human Neuropixels workflow outside of the US, several limitations must be acknowledged. First, the small sample size (n=6) reflects the early developmental nature of the project. While this cohort is sufficient to monitor the procedural learning curve and identify failure modes, larger studies are required to establish definitive benchmarks for unit yield and characteristics across different hospital environments and research paradigms. Consequently, we do not present biological analysis of the acquired neural data. The viable recordings obtained during this early developmental phase are insufficient in number to support robust mechanistic conclusions. Further, single-unit analysis is inherently more reliable when contextualised within a larger, standardised cohort. Therefore, the scope of this manuscript is intentionally restricted to methodological and regulatory validation. Second, our focus on participants under GA, necessitated by our frameless insertion methodology, limits the ability to perform awake, task-based paradigms. The anaesthetised brain offers a unique window into fundamental neurophysiology, and future recordings may combine non-invasive sensory stimulation. Finally, the technical outcomes of this study highlight the inherent challenges of IDEAL 2a development. While the Neuropixels probe was successfully inserted and explanted without clinical complication in all six participants, we encountered technical barriers that precluded successful spike sorting in 50% of the cohort. In two cases, significant intraoperative electromagnetic noise, subsequently identified as the neuronavigation system, compromised the signal such that no usable data could be collected. In a third case, the recording duration was insufficient to allow for robust motion correction and automated spike sorting. Rather than viewing these as failures, these cases served as vital data points that facilitated the iterative development of theatre de-noising steps in the workflow to increase overall reliability.

## Conclusion

The translation of Neuropixels from a preclinical tool to a clinical research instrument requires a systems-engineering approach that prioritises safety, efficiency, and reproducibility. Using the IDEAL Stage 2a framework, we demonstrate that a standardised intraoperative Neuropixels workflow can be successfully integrated within a centralised healthcare and regulatory environment.

Our study achieved its primary safety endpoint (E1), with no shank fractures or research-related adverse events observed. High-density recordings were consistently performed within a capped 30-minute procedural extension (E2), supporting integration into routine elective surgical workflows. Iterative refinement of the technical and procedural workflow resulted in progressive improvements in data fidelity (E3), culminating in a stabilised configuration yielding 146 putative units within a 15-minute recording window.

This workflow’s use of manufacturer-sterilised hardware and a frameless insertion methodology reduces key technical barriers to human Neuropixels recordings and demonstrates feasibility beyond specialised awake-mapping environments. Future work aligned with IDEAL Stage 2b (Exploration) will focus on multi-centre validation and benchmarking of neural data quality. This framework provides a foundation for scalable and reproducible human single-unit electrophysiology, supporting future integration with multimodal human neuroscience research.

## Data Availability

This paper does not report datasets deposited in a public repository. De-identified neural recordings supporting the findings of this study are not publicly available as analysis is ongoing, but are available from the lead contact upon reasonable request, subject to ethical approvals and a data access agreement. The pre-surgery checklist is provided as Appendix 1 in the supplemental information. This paper does not report original code.

## Acknowledgments

We would like to thank the National Hospital for Neurology and Neurosurgery anaesthetic and theatre teams (Dr Val Luoma and Immaculate Ignacio) and UCLH Biomedical Engineering (Thomas Hague, Alyte Podvoiskis, Mike Notaridis). We would also like to thank the UCL Comprehensive Clinical Trials Unit (Grace Auld, Rachel McComish, Connor McAlpine, Atiyyah Moosa), Bart de Strooper and Lorena Carcamo at the Francis Crick Institute for the scientific discussions, and Leigh Hochberg at Massachusetts General Hospital / Brown for helping to launch the collaborative research.

## Author Contributions

HLH, ATS, MK and WM conceptualised the study, designed the methodology, and wrote the original draft. HLH, MK, WM, AKT, and LW conducted the clinical investigation. HLH, LW, AKT, WM contributed to participant recruitment. BFC, ACP and SSC shared foundational methodology. AS, JC, AV designed and fabricated the phantom used for simulation. MW and BD provided sterilised Neuropixels hardware and expert advice. ATS provided supervision, funding acquisition, and resources. MK performed formal data analysis, including spike sorting and manual curation. All authors contributed to review and editing.

## Methods

| REAGENT or RESOURCE | SOURCE | IDENTIFIER |
| --- | --- | --- |
| **Deposited Data** |  |  |
| Pre-operative surgical checklist | This paper |  |
| **Software** |  |  |
| SpikeGLX |  |  |
| OpenEphys GUI | Open Ephys |  |
| Kilosort 4.0 | Mouseland |  |
| MEDiCINE (Motion Correction) |  |  |
| **Hardware** |  |  |
| Neuropixels 1.0 NHP short-S probes | Imec |  |
| Headstage (HS_1000) | Imec |  |
| Cable (CBL_1000) | Imec |  |
| Metal cap rod holders | Imec |  |
| Onebox acquisition system | Imec |  |
| Vexio-Cart acquisition stack | Parity Medical |  |
| Dual Elo 2403lm medical-grade monitors | Elo |  |
| Endoscope holder | Karl Storz |  |
| Universal clamping jaw | Karl Storz |  |
| Rotation socket | Karl Storz |  |
| Mitaka micromanipulator | Mitaka |  |
| Point setter universal adapter | Mitaka |  |
| Custom sterilisation tray | RB medical |  |
| Subdermal needles (Grounding) | Medtronic | 945dsn2280 |
| **Chemicals** |  |  |
| Ecoflex GEL (Part A) | Smooth-On, Inc. | FG-823A |
| Ecoflex GEL (Part B) | Smooth-On, Inc | FG-823B |
| Talcum powder | Johnson & Johnson | EAN: 3574669908252 |
| MED-1000 | Polymer Systems Technology Ltd. | NuSil ID: MED-1000 |
| n-heptane | Sigma-Aldrich | Cat#592579; CAS 142-82-5 |

## Resource availability

### Lead contact

Requests for further information or resources should be directed to and will be fulfilled by the lead contact William Muirhead or Hugo Layard Horsfall.

### Materials availability

This study did not generate new unique reagents.

### Experimental model and subject details

Six participants were prospectively recruited at the National Hospital for Neurology and Neurosurgery, a quaternary academic neurosciences centre (n=6; 1 male, 5 female; mean age 62.5 years; range 41-79 years). Diagnoses included idiopathic normal pressure hydrocephalus (n=3), late midlife hydrocephalus (n=2), and early midlife hydrocephalus (n=1). All recordings were performed in the right parieto-occipital cortex (Brodmann area 39; angular gyrus) during elective ventriculoperitoneal (VP) shunt insertion.

One participant (SU004) was excluded from analysis as the procedure was postponed post-consent due to clinical comorbidities. The maximum extension to the planned operation was 30 minutes, of which a maximum of 15 minutes could be used for Neuropixels recording. All participants were under general anaesthesia (Table 1).

Inclusion criteria:

1. Aged 18 years and over
2. Documented informed consent to participate
3. Undergoing a transcranial neurosurgical procedure at the National Hospital for Neurology and Neurosurgery (NHNN) in which part of the cerebral cortex will be:

a. Resected
b. Transgressed as part of the surgical approach to reach a deeper target
c. Transgressed by the passage of a surgical implant
4. Surgery technically suitable

Exclusion criteria:

1. Participants who lack the capacity to give informed consent
2. Participants who are pregnant or breast feeding at the time of surgery
3. Participants deemed to be unsuitable for recruitment to the study by the responsible Consultant Neurosurgeon
4. Concern from the Consultant Neurosurgeon or treating Anaesthetist that there would be an unreasonable burden on the patient to participate in the study
5. For participants requiring anaesthesia for their pre-planned brain surgery, an American Society of Anaesthesiologists (ASA) physical status classification system score of 4 and higher
6. Participants unable to understand English sufficiently well to read the patient-facing documents and consent in English.
7. Participants not undergoing their treatment through NHS care (i.e. private patients are excluded).

## METHOD DETAILS

### Regulatory and ethical approvals

This study was conducted in accordance with the principles of the Declaration of Helsinki and Good Clinical Practice (GCP). Full ethical approval was granted by the Health Research Authority (HRA) and the regional Research Ethics Committee (REC reference: 24/NE/0226), with the University College London Comprehensive Clinical Trials Unit (UCL CCTU) acting as the study sponsor. The study is registered with ISRCTN (ISRCTN25803759) and ClinicalTrials.gov (NCT06763471).

As detailed in the Results section, the project was established as a non-commercial basic science study with human participants. Following formal consultation with the Medicines and Healthcare products Regulatory Agency (MHRA), the Neuropixels probe was legally classified as a research tool, exempting the study from formal clinical investigation notification under the UK Medical Device Regulations (UK MDR).

All participants provided informed written consent prior to inclusion. To ensure robust ethical safeguarding, a mandatory 24-hour cooling-off period was enforced between the provision of the patient information sheet and the signing of the consent form.

### IDEAL Stage 2a study design and evaluation metrics

We prospectively validated our human Neuropixels workflow using an IDEAL Stage 2a (Development) framework(McCulloch et al., 2009) to monitor the learning curve and iteratively refine the surgical and technical workflow through a transparent change history. Development was evaluated against three primary clinical and technical endpoints:

- Endpoint 1 (E1): safety (Notifiable Safety Events; NSE). This included the incidence of Neuropixels shank fractures and other research-related adverse events (haematoma, infection, or wound complication) monitored intraoperatively and at 30-day follow-up.
- Endpoint 2 (E2): duration. We recorded the total research operative extension time (capped at 30 minutes) and the Neuropixels recording time (maximum 15 minutes).
- Endpoint 3 (E3): data. This was defined by the binary acquisition of neural data (Yes/No) and the subsequent yield of putative units following automated spike sorting.

Workflow stabilisation was tracked through iterative versions (v0.1 to v2.0), with process changes triggered by documented failure modes and verified in subsequent clinical cases. Maturation was driven by a systematic logic whereby technical or environmental challenges were identified as failure modes, analysed for root causes (e.g., noise from neuronavigation), and addressed through specific mitigations or workflow iterations. Each iteration was verified in the subsequent clinical case to confirm technical maturation and assess residual risk. Safety oversight included a pilot phase at n=5 for initial safety review by the trial steering committee. Through pre-clinical simulation and using a silicone brain phantom that, that should probe fracture occur, a probe retrieval protocol was established using shunt-protected toothed forceps which enabled dextrous grabbing of the shank without causing it to shatter.

### Phantom fabrication

Base layer preparation: The base layer consisted of a silicone-based gel and talcum powder, combined in a 1:1:0.477 weight ratio of Ecoflex GEL parts A and B to talcum powder. Both parts of Ecoflex GEL were stirred thoroughly within their storage containers, then poured into a plastic cup and mixed for 30 s, prior to the addition of talcum powder, to ensure a successful curing process. Once added, the mixture with talcum powder was stirred for three minutes before degassing at 100 kPa until entrapped air had been removed, then left to cure at room temperature for two hours. Following curing, excess plastic from the cup was trimmed flush with the phantom base.

Top layer preparation: MED-1000 was mixed with n-heptane in a 1:1 ratio to reduce the viscosity of MED-1000 which enabled it to be used for spin coating without disturbing the curing process. 5 ml of each of MED-1000 and n-heptane were loaded into two 10 ml syringes. The two syringes were then connected with a Leur-Lock syringe cap to mix the contents by pushing the syringes back and forth 50 times (Schuettler et al., 2005). Extra precaution was taken to ensure no excess air was caught in either syringe. A flat platen (9cm diameter) was covered with adhesive tape (TESA 4124), sprayed with Isopropyl Alcohol (IPA) and wiped dry. Two millilitres of the MED-1000+heptane mixture was deposited onto the centre of the platen whilst spinning at 100 RPM for 15 s with a 20 RPM ramp, followed by 400 RPM for an additional 90 s once the mixture was fully applied. Once complete, the tape was removed and placed on a baking sheet to cure in an oven at 68° C for an hour.

Phantom assembly: Once removed from the oven, the MED-1000 film was peeled off the tape and placed onto the base layer by stretching it evenly from all four corners until taut. It was uniformly lowered onto the base to minimise trapped air, sticking to both the plastic cup and base layer without any additional adhesives. The phantom may be stored at room temperature with negligible degradation.

### Neuropixels hardware and sterilisation

Neuropixels 1.0 NHP short-S 10 mm long probes with a thickened 122 µm shank and sharpened tip (Imec, Belgium) were used for all recordings. This design represents an evolution from earlier 24 µm shanks to reduce pial penetration resistance and negate the requirement for intraoperative piotomy (Chung et al., 2022; Paulk et al., 2022). In contrast to previous protocols requiring in-house assembly and soldering by researchers, our workflow utilised manufacturer-led (Imec, Belgium) end-to-end sterilisation via Ethylene Oxide (EtO). All components, including the probe, headstage (HS_1000+CBL_1000-S), and metal cap probe holders (HOLDER_1000_C-S), were delivered in surgical-grade blister packs or pouches with pre-soldered grounding and referencing leads, reducing technical friction in the sterile field.

### Neuropixels preparation and functional testing

To maintain sterility and clinical integrity, initial preparation was conducted in the theatre scrub room. Sterile equipment was prepared on a sterile-draped large trolley. A sterile kidney dish was half-filled with warm 0.9% sodium chloride. Once draping of the patient had commenced, the research team moved into the operating theatre to begin formal setup. The Neuropixels headstage was attached to the probe whilst in its protective blister pack. Once connected to the acquisition stack and headstage tests complete, a mosquito clip was used to gently remove the probe via the metal cap for mounting onto the imec rod holder. Two narrow steristrips were applied to the metal cap to create handling wings (Paulk et al., 2022). The distal end of the rod holder was secured to the sterile kidney dish using mosquito clips and steristrips. The rod was inverted and stabilised by an upside-down saline jug, allowing the probe to be half-immersed in saline. Subdermal needles (Medtronic, 945dsn2280) were connected to the pre-soldered grounding leads and placed in the saline to complete the circuit. A comprehensive diagnostic suite was run via SpikeGLX and/or OpenEphys to confirm the integrity of the recording setup. Baseline noise was checked to ensure signal stability prior to transfer to the operative field. This stage did not contribute to research time (E2: duration) as the planned operation was underway in parallel.

### Surgical suite and frameless insertion

Participants were positioned supine in a standard horseshoe headrest without head-fixation in a Mayfield clamp. The insertion equipment used an endoscope holder, universal clamping jaw, and table clamp (Karl Storz, Germany) connected to a micromanipulator and point setter universal adapter (Mitaka, Japan). This configuration provided macroscopic alignment with multiple degrees of freedom and sub-millimetre precision in pitch, yaw, and translational advancement. A manufacturer-sterilised rod holder (Imec, Belgium) was used to mount the probe to facilitate orthogonal and stable insertion into the cortex (Chung et al., 2022).

### Preparation for recording and intraoperative de-noising protocol

When the consultant neurosurgeon was at an appropriate operative stage and ready to commence the research component, a checklist systematically de-noised the operative environment. Once verbal feedback confirmed the equipment was managed, two study surgeons (W.M and H.L.H) transferred the endoscope holder pre-attached to the micromanipulator to the operative field and grossly aligned it to the region planned for Neuropixels insertion. The dura is then opened in line with routine VP shunt procedures. The rod holder with Neuropixels and grounding cables is transferred to the operative field, attached to micromanipulator and subdermal needles placed in the scalp separately and as far as reasonably possible within the confinements of the operative field as outlined by Paulk et al., 2022. Care must be taken when handling the Neuropixels as it is extremely fragile and we recommend a “pincer grip” (Figure 1l) (Coughlin et al., 2023). Small adjustments of the endoscope holder and micromanipulator optimise perpendicular trajectory.

### Neuropixels recording

Due to the nature of the planned operation, ventriculoperitoneal shunt insertion, burr holes are performed at homogenous sites. This limits the exposed cortex compared to a large craniotomy, and the Neuropixels must be inserted in the same region for planned ventricular catheter insertion. Care is taken to avoid large vessels on the cortical surface. Verbal communication between two surgeons (W.M and H.L.H) to achieve as close to orthogonal insertion trajectory using the submillimetre control of the micromanipulator. The Neuropixels is then driven using the micromanipulator and inserted through intact pia without additional piotomy, towards a target depth of 5-7mm. The 15-minute recording window begins once the Neuropixels is in cortex. Following the recording, or if the recording was aborted due to noise, the Neuropixels was retracted at similar speed to insertion using the micromanipulator. After explantation, the cortical surface was visually inspected for evidence of bleeding. The Neuropixels was protected to avoid shank fracture and the subdermal needles removed and placed in sharps bin. The Neuropixels en bloc was transferred back to the preparation table and a photo of the shank was taken to ensure it was intact. The operation then continued as planned.

## QUANTIFICATION AND STATISTICAL ANALYSIS

### Neural data acquisition and sorting

The acquisition stack was a Parity Medical Vexio-Cart equipped with a 660VA isolating transformer and dual Elo 2403LM medical-grade monitors. Initial cases utilised a National Instruments PXIe chassis (NI PXIe-1071; NI PXIe-6341; Neuropixels PXIe Acquisition Module); following hardware instability in SU006, the imec Onebox acquisition system was integrated for subsequent cases. Neural data were acquired via OpenEphys GUI at a sampling rate of 30 kHz for the action potential (AP) band and 2.5 kHz for the local field potential (LFP) band, with hardware gains of 500× (AP) and 250× (LFP) in accordance with standardised Neuropixels 1.0 protocols.

Post-hoc spike sorting was performed using Kilosort 4.0 with motion correction via MEDiCINe. From 761 candidate units, manual curation was performed by an experienced neuroscientist (M.K.) using SpikeInterface GUI, based on waveform morphology, inter-spike interval violations, and signal-to-noise ratio, and yielded 146 single units (172 multi-unit activity, 443 noise). Quality metrics were computed using SpikeInterface (firing rate: 1.35 ± 3.35 Hz; SNR: 6.6 ± 6.5; spike amplitude: 262 ± 242 µV; ISI violation rate: 0.29 ± 0.94%; presence ratio: 0.57 ± 0.37). Detailed commentary on spike sorting methodology and neural data analysis is beyond the scope of this paper.

**Supplementary Figure 1.**
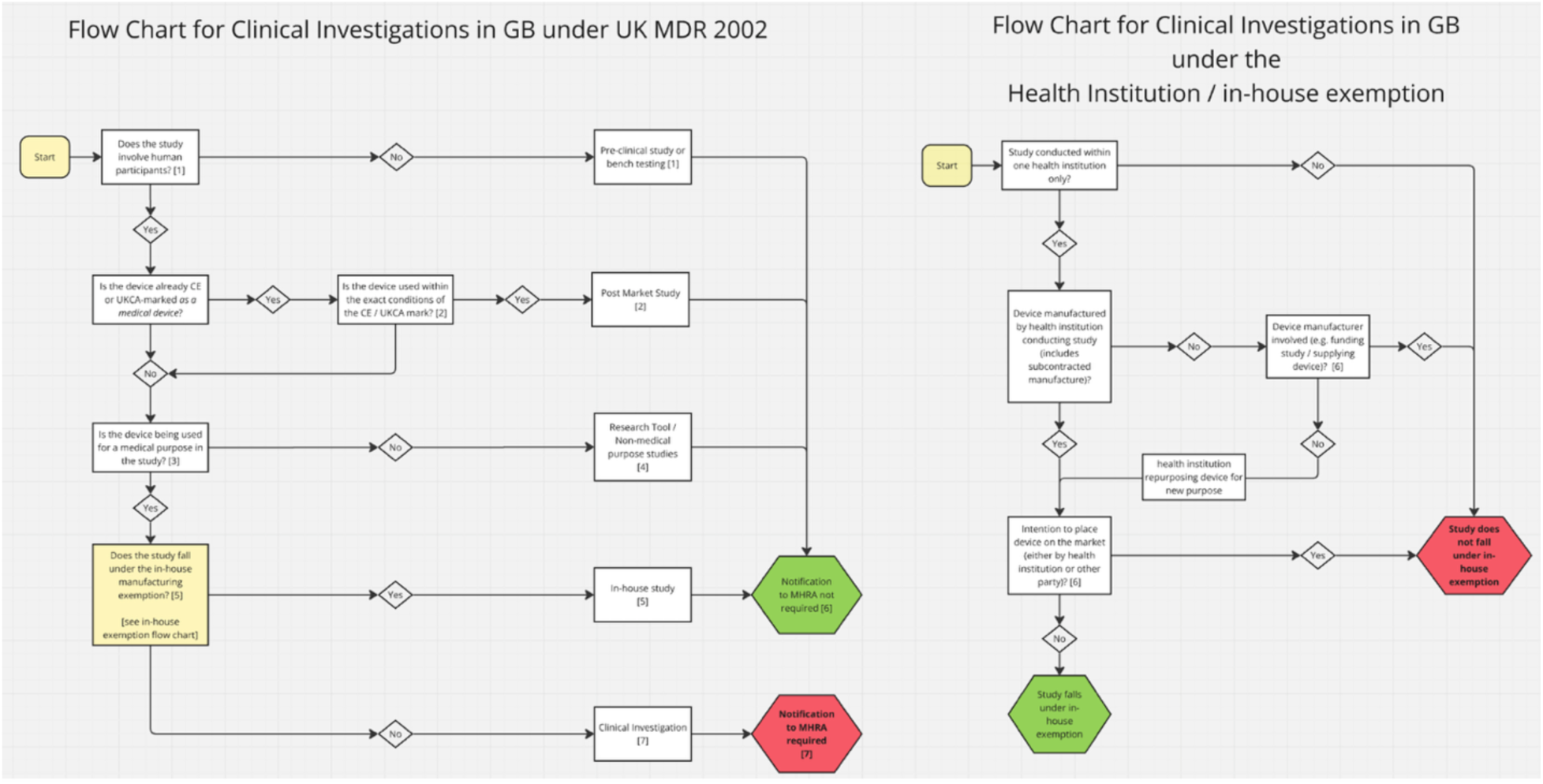
Flowchart for regulatory decision making. Medicines and Healthcare products Regulatory Agency online Flow Chart for Clinical Investigations under UK Medical Device Regulations (https://assets.publishing.service.gov.uk/media/69247b5dba812a67cb6a569d/MHRA_Flow_Chart_for_Clinical_Investigations_Under_UK_MDR_Version3.png; https://www.gov.uk/guidance/notify-mhra-about-a-clinical-investigation-for-a-medical-device; accessed 01 April 2026).

**Supplementary Figure 2.**
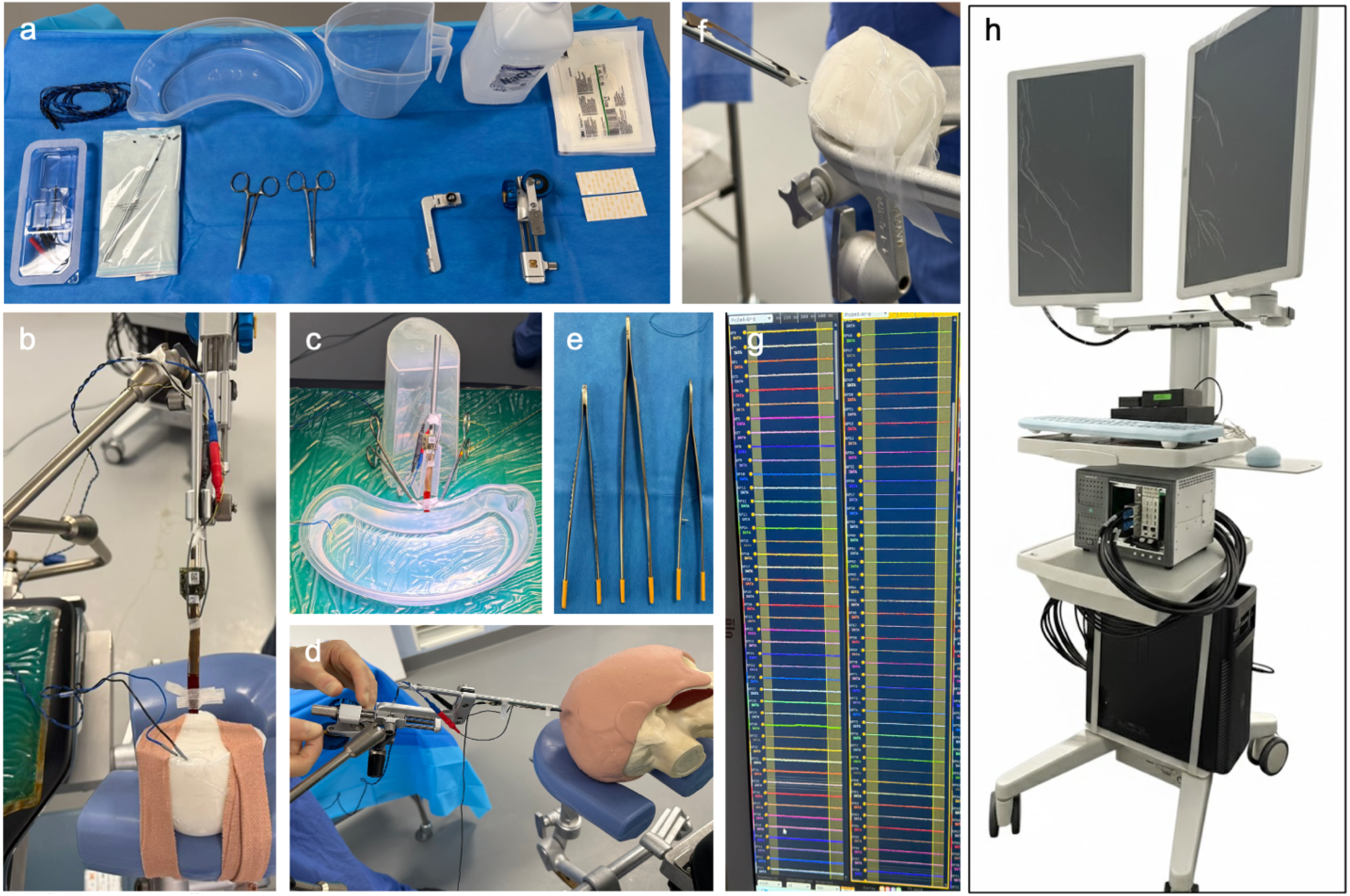
Pre-clinical simulation, electrical de-noising and fracture retrieval. **(a)** Sterile equipment setup as on day of surgery. Equipment left to right: sterilised Neuropixels in blister pack, subdermal needles for grounding, rod holder, kidney dish for sterile saline, jug, mosquito clips, micromanipulator with attachment, steristrips. **(b)** Simulation of pial penetration using bespoke brain phantom. Karl Storz endoscope holder attaches the micromanipulator to the operating table and provides multiple degrees of freedom. **(c)** Baseline electrical noise testing on operating table with complete circuit. Theatre lights turned to maximum brightness and shone directly onto Neuropixels to assess noise. **(d)** Simulated inserted with skull phantom for ventriculoperitoneal shunt surgery with frameless technique and horseshoe headrest. **(e)** Toothed forceps protected with shunt tubing for shank fracture retrieval to prevent shank shattering. **(f)** Verification of shank retrieval without shattering using protected tooth forceps. **(g)** Example background noise from operating theatre on OpenPhys.

**Supplementary Figure 3.**
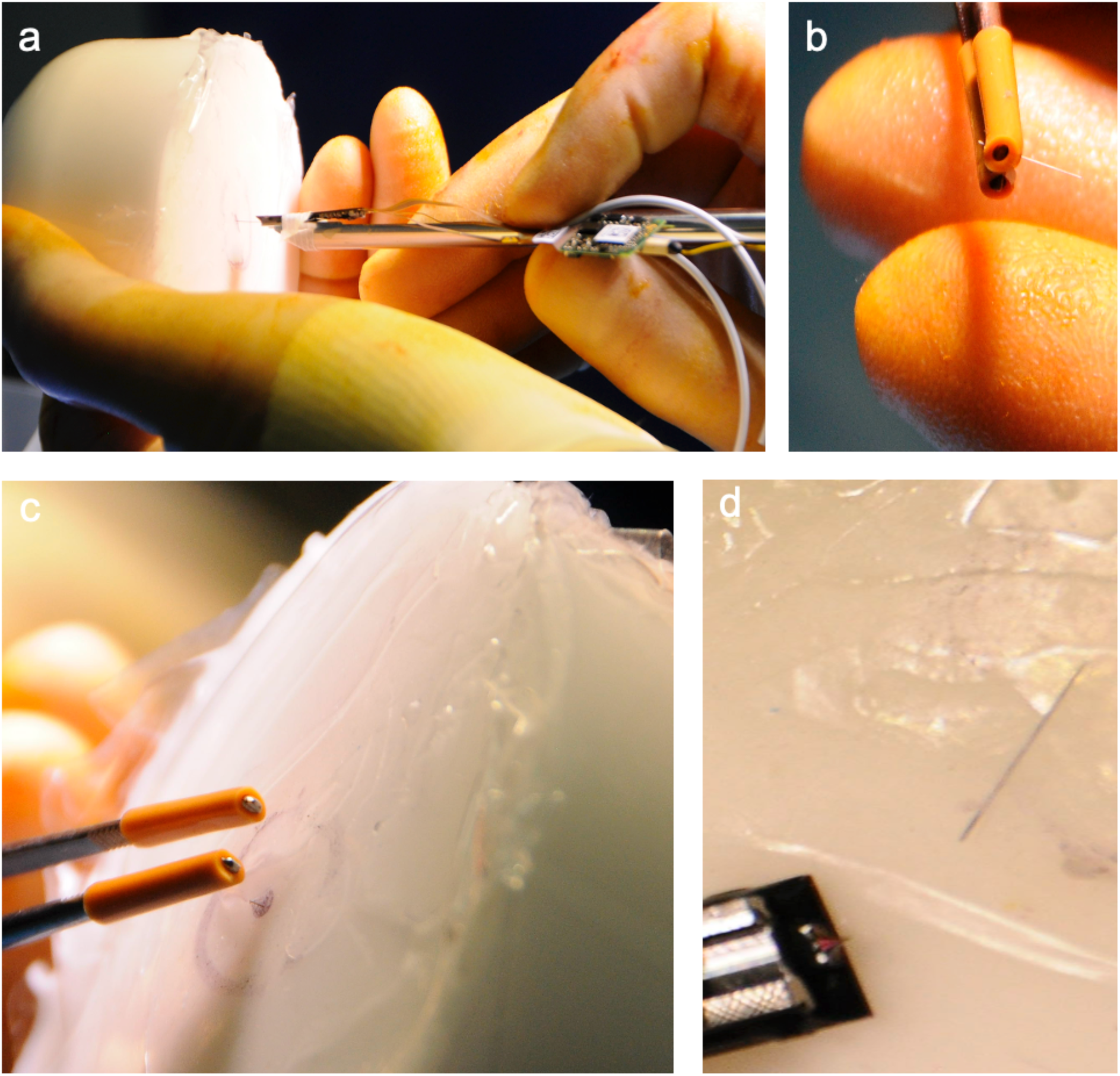
Shank fracture and successful retrieval during simulation. **(a)** Simulating mechanical failure mode during insertion resulting in shank fracture in brain phantom (see Methods). **(b)** Using shunt tubing protected tooth forceps to prevent shank shattering, shank is removed en bloc successfully. **(c)** Difficult shank fracture retrieval with majority of shank in the “cortex”. Success demonstrated in previous panel. **(d)** Confirmation of mechanical fracture occurs at proximal end of shank in agreement with literature: Paulk et al 2022, Chung et al 2022, Chung et al 2025.

## Appendix 1. Pre-surgery checklist

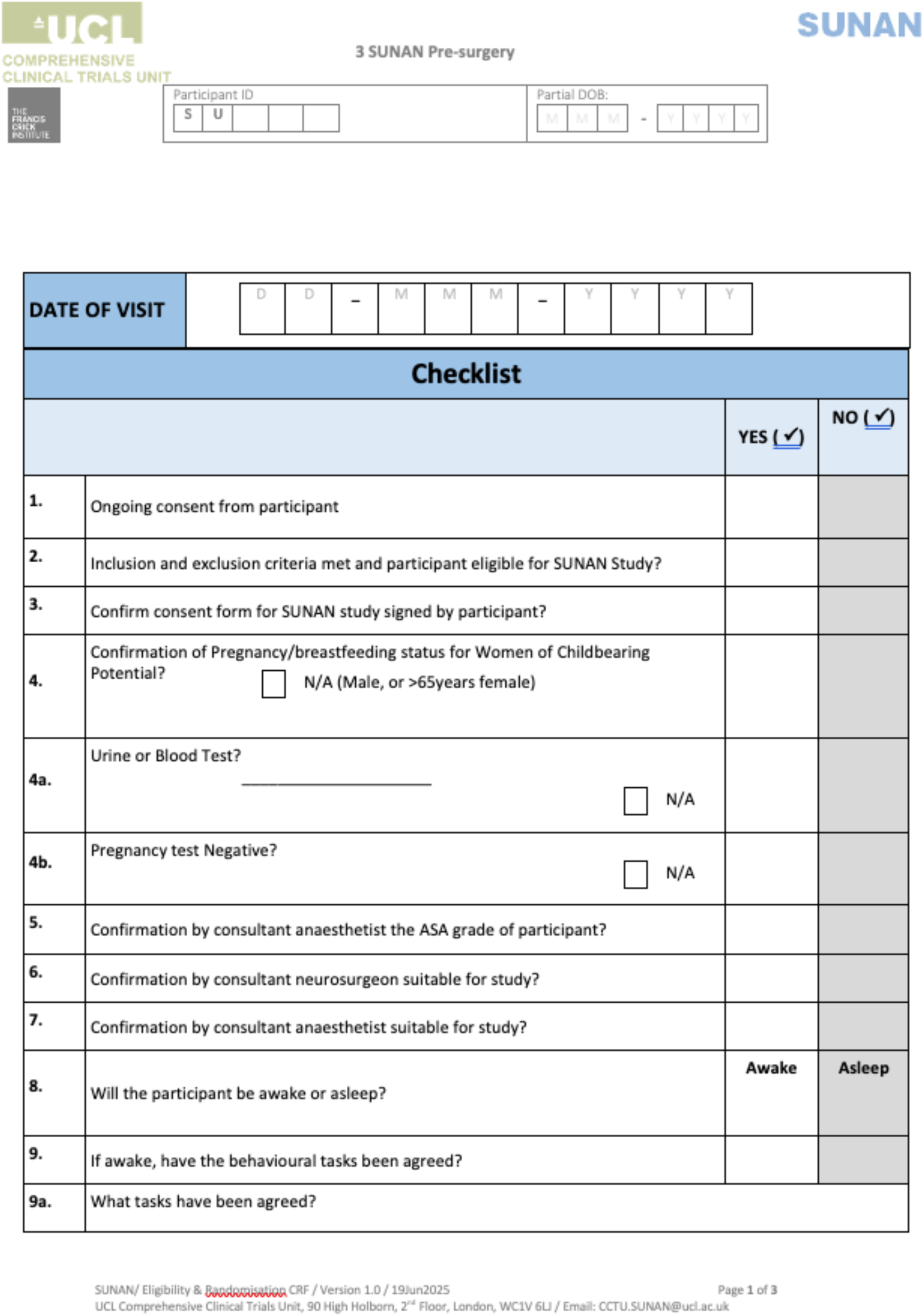
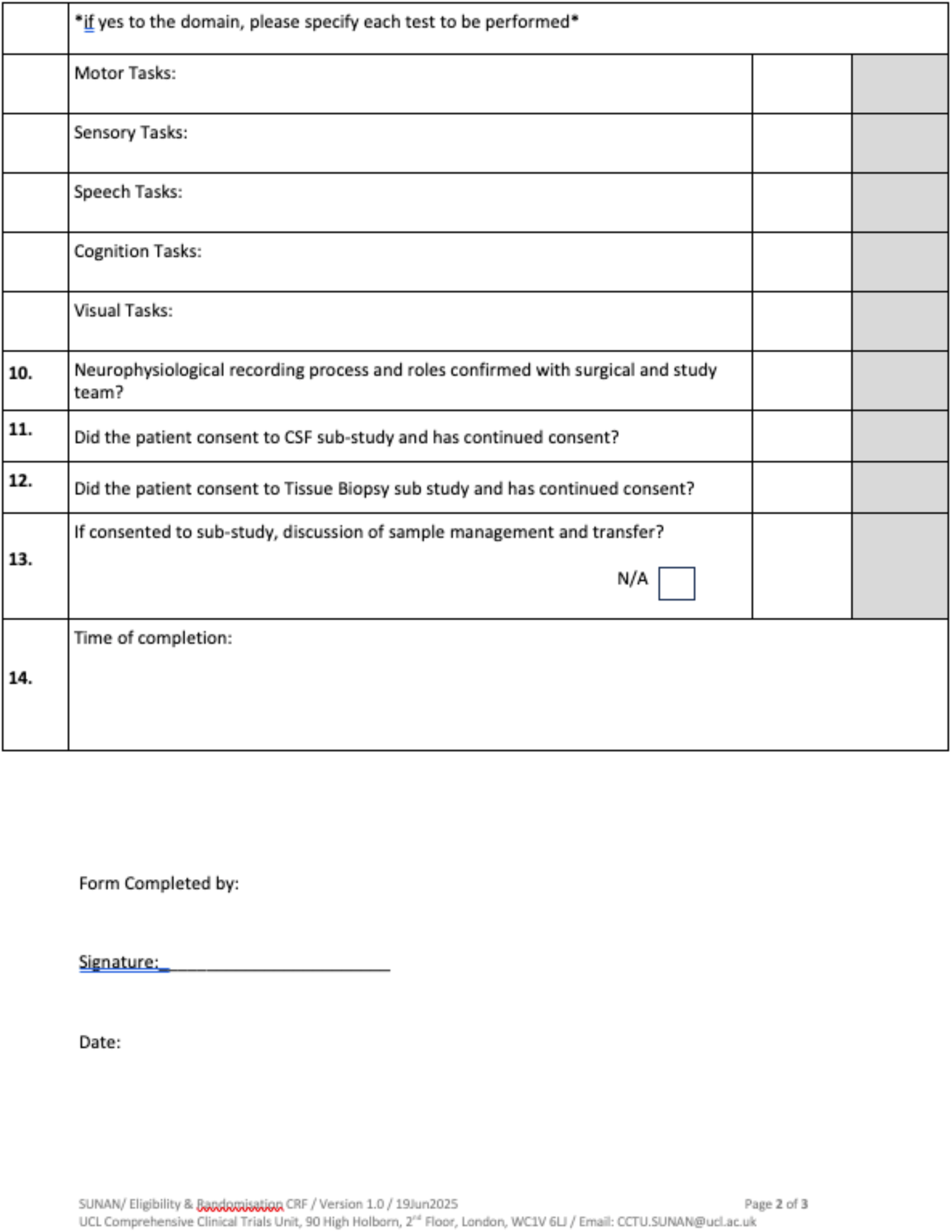

**Supplementary Table 1.**
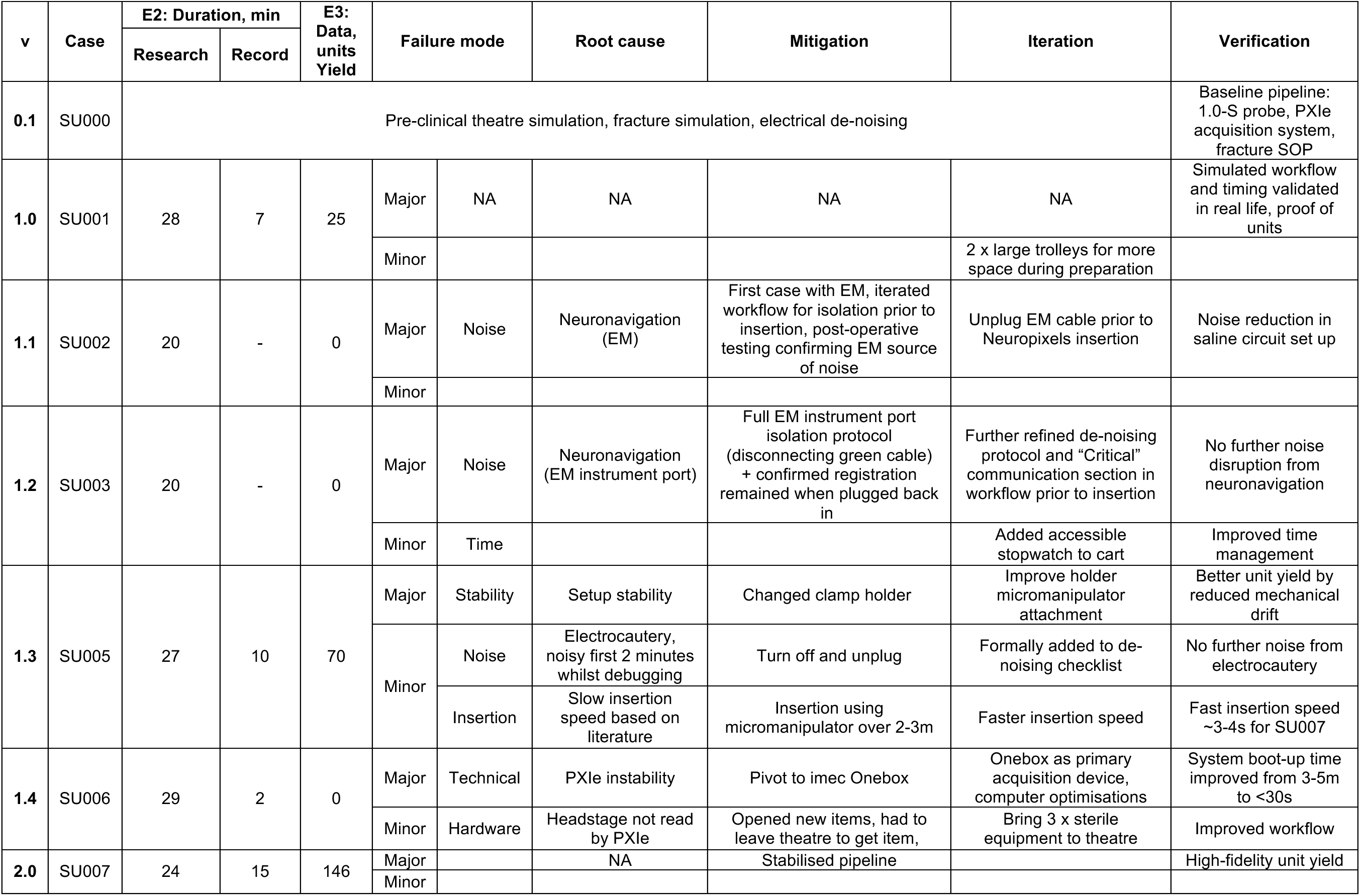
Iterative technical and procedural refinements during IDEAL Stage 2a Development. Prospective documentation of iterative pipeline maturation across consecutive surgical cases using the IDEAL Stage 2a Development framework. Each version reflects modifications introduced in response to observed failure modes with key Endpoints (E) of Duration and Data. Notifiable Safety Events were not included in this table as there were none. Failure mode was categorised as Major or Minor depending on impact on E2 and E3. Mitigation strategies were implemented and validated in subsequent cases. Recording duration and data yield improved iteratively, culminating in a stabilised pipeline (v2.0) achieving 146 putative units during the full 15-minute recording window. These iterative refinements informed the development of standardised workflows, intraoperative de-noising protocols, and integration of hospital-compatible acquisition hardware. Further pre-operative and post-operative iterations relating to recruitment, consent, data management, and general study iterations are not considered in this table. v: Version of pipeline; E2: Endpoint 2; E3: Endpoint; EM: electromagnetic neuronavigation

